# Black Women’s Maternal Health: Insights from Community Based Participatory Research in Newark, New Jersey

**DOI:** 10.1101/2023.03.10.23287120

**Authors:** Leslie M. Kantor, Naomi Cruz, Christiana Adams, Constance Akhimien, Fauziyya Allibay Abdulkadir, Cherriece Battle, Maria Oluwayemi, Olanike Salimon, Sang Hee Won, Teri Lassiter, Sophee Niraula

## Abstract

This study on Black women’s maternal health engaged a group of six community members in a community based participatory research project in a state with one of the largest racial disparities in maternal mortality and severe maternal morbidity in the United States. The community members conducted 31 semi-structured interviews with other Black women who had given birth within the past 3 years. Four main themes emerged: (1) challenges related to the structure of healthcare, including insurance gaps, long wait times, lack of co-location of services, and financial challenges for both insured and uninsured people; (2) negative experiences with healthcare providers, including dismissal of concerns, lack of listening, and missed opportunities for relationship building; (3) preference for racial concordance with providers and experiences with discrimination across multiple dimensions; and (4) mental health concerns and lack of social support. CBPR is a research methodology that could be more widely deployed to illuminate the experiences of community members in order to develop solutions to complex problems. The results indicate that Black women’s maternal health will benefit from multi-level interventions with changes driven by insights from Black women.

## Introduction

The United States (US) is the only high-income country in which maternal mortality has increased over the past decade,^1^ and racial disparities in maternal health are stark. In 2020, the maternal mortality rate nationally for non-Hispanic Black women was more than double that of White women at 55.3 deaths per 100,000 live births, compared to 19.1 deaths per 100,000 live births for non-Hispanic White women.^2,3^ Black women are also significantly more likely to die or experience severe maternal morbidity (SMM) than White women.^4,5^ States vary considerably in their maternal health indicators, and New Jersey has among the highest rates of maternal mortality in the US at 24.1 deaths per 100,000 live births.^6^ New Jersey also exhibits marked racial disparities in maternal mortality, with 79.6 deaths per 100,000 live births for Black women versus 16.8 deaths per 100,000 live births for White women.^7^

In addition to negative clinical outcomes, reports of disrespect, mistreatment, and other negative patient experiences both internationally and domestically have raised calls to define and implement respectful maternity care.^8^ A number of qualitative investigations have recently been undertaken to better understand Black women’s reproductive and maternal health experiences in the US and identify opportunities for improvement. ^9–13^ These studies utilize a variety of methods that partner researchers with community-based organizations (CBOs) and community members in various ways.^9–13^ Similar research has been conducted outside of the US with other populations that experience maternal health disparities, such as migrant women.^14,15^ Community-engaged approaches to research are critical to ensuring that the lived experiences of Black women are accurately reflected, and community-based participatory research (CBPR) is a particularly salient approach to better understanding opportunities for improvements throughout the prenatal, labor and delivery and post-partum phases of pregnancy.

CBPR is fundamentally rooted in the ideals of equity and community empowerment.^16,17^ The basic premises of CBPR are reflected in nine principles meant to ensure the equitable and active involvement of community members throughout all stages of the research process.^16^ As Wallerstein and Duran note: “Community-based participatory research (CBPR) has emerged in the last decades as a transformative research paradigm that bridges the gap between science and practice through community engagement and social action to increase health equity.”^18^ This is demonstrated by studies which suggest that greater levels of participant trust and engagement may stem from employing community-engaged methods which can bolster the overall acceptance and effectiveness of subsequent interventions.^19–22^ CBPR has the potential to glean insights that will help reduce health disparities.^23–25^

Recent studies on Black women’s maternal health utilizing a CBPR framework or approach have found that Black women frequently felt that their concerns were dismissed; their interactions with healthcare providers and staff were stressful; they had unmet needs for information; and they lacked social support.^10,13^ Many of these investigations have been undertaken in particular geographic contexts; however, other areas with maternal health disparities offer additional insights. Thus, this investigation was undertaken in Newark, New Jersey which has one of the highest rates of maternal mortality and largest disparities between Black and White women in the country.

The study was approved by the university IRB: Pro2019002981.

## Methods

This study used CBPR methods to engage Black mothers as equitable research partners in all phases of the present study. Recruitment for the community researchers was conducted by our CBO partner which enlisted its network of organizational partners in promoting the opportunity. A total of six Black mothers were recruited and trained as community researchers to center the experiences of pregnancy, labor, delivery, and postpartum among Black women. All of the Black mothers completed a four-session training on research techniques, including training in the protection of human subjects. The full research team met 16 times over the course of the study to plan participant outreach, debrief about interviews, analyze the data, and create recommendations.

Black women who gave birth within the last three years and resided in Newark area were recruited by community researchers through contact with Greater Newark Healthcare Coalition. Potential participants were directed to the Rutgers School of Public Health’s online research system, which included a brief video of one of the community researchers explaining the study, a screening questionnaire, and consent forms. Interviews were scheduled by a university research assistant and conducted remotely, using Zoom. The remote interviews took place between January-July 2022 and ranged from 19-70 minutes. Participants received $50 gift cards for completing the interviews. Interviews were transcribed by a professional transcription company after a quality review of Zoom transcripts found multiple errors.

Data analysis was conducted using a general inductive approach to find and clarify patterns and themes in the data.^26^ The full research team worked together initially to analyze transcripts and identify codes and then utilized the codes in reviewing transcripts. Each transcript was carefully reviewed by one community researcher and one university researcher and each reader also created a brief narrative summary of the main issues in the transcript and key participant quotes. Following this process, the team reviewed codes that were closely related and collapsed them into themes. The process was designed to be as accessible as possible to the community researchers, consistent with recent thinking on adapting qualitative methods to achieve health equity.^27^

## Results

Forty-seven people were screened and eligible for the study, and 31 interviews were completed. The 16 eligible participants who did not complete the interviews did not respond to multiple attempts to schedule or did not attend the scheduled interview time despite additional follow-up attempts. There were a wide range of experiences reported throughout the pre-natal, labor, delivery, and post-partum phases of pregnancy. Four major themes emerged across the 31 interviews: (1) structural challenges to obtaining healthcare (2) negative experiences with healthcare providers including dismissal of concerns, lack of listening, and missed opportunities for relationship building; (3) preference for racial concordance with providers and reports of discrimination based on age, weight, and type of insurance; and (4) mental health concerns and lack of social support.

### Structural Challenges to Obtaining Healthcare

Analysis of the interviews illuminated key structural challenges that interfered with pregnant women’s abilities to obtain healthcare, including gaps in insurance coverage, financial challenges for both insured and uninsured people, long wait times for short appointments, and lack of co-location of services, which was particularly problematic for people who relied on public transportation or did not have paid sick time.

While participants in the study had a range of insurance coverage, many, especially new immigrants, faced challenges obtaining insurance during pregnancy which led to delays in care. One participant observed: “Whether you are legal or illegal…they should not discriminate when it comes to health insurance” (P15). Another participant shared: “I didn’t have health insurance during my first trimester because I was trying to get it, and they were sending me back and forth. So, all through the first three months, I wasn’t checked” (P08). Even for people with insurance coverage, many still faced challenges paying for the costs of services and/or medications: “I had to stretch out my appointments to make sure that I had enough funds to cover the copays” (P27).

Wait times for medical appointments and labs at multiple locations were other structural challenges that emerged from the interviews. Participants offered their perspectives on the causes of these structural problems, noting that healthcare providers and hospitals were understaffed and overscheduled. As one participant noted: “I did feel like the visits were kinda rushed. I didn’t get a lot of time with [the doctor] in the room. Most of the time I was with the medical assistant” (P14). Many participants also experienced difficulty obtaining lab services and procedures because of the complexity of getting to multiple locations. Several participants described challenges in paying for recommended supplements such as pre-natal vitamins that were not covered by insurance and wanted providers to prescribe these medications to have them covered and to ensure they were taking exactly what was being described.

See Table 1 for additional quotes about structural challenges.

**Table 1.** Additional Representative Quotes for Structural Challenges to Obtaining Healthcare.

|  |  |
| --- | --- |
| “I don’t really understand why it’s not easy for someone who’s pregnant to just, you know, for them to get insurance easier...In other states, like I’ve lived in Florida before, and once you’re pregnant, you just apply online and you put that you are pregnant, they automatically give you Medicaid...But I felt like, over here, it’s like you have to bring this and bring that and you have to search for it.” | P02 |
| “Yeah, my insurance ended while I was on maternity leave. And I was like, ‘why does that happen?’ I work for ***; it shouldn’t happen like that. They shouldn’t make it so hard for me when I’m on maternity leave.” | P10 |
| “They had some stuff that really—I was not really happy about it because I was pregnant, and I told her like ‘I have to see the doctor,’ you know, as I’m pregnant. And she could see that my tummy was [inaudible] showing, and she was like, ‘No, no, no you have to do this except you go to the emergency and you pay for it.’...So that really got me very sad, and I just left the office, and I went back until [until I was approved] and that was quite late. Even the doctor was surprised why I didn’t come to the hospital.” | P06 |
| “I was going [to the doctor] three times [a week]. My insurance is not paying for that. Up till now, we’re still trying to sort it out. It’s been two years already.... So, they are not paying for those tests. That’s a lot of money.” | P19 |
| “I would say with the billing because they checked me as an international patient...Yeah, it was quite expensive, because my whole hospital bill was over \$10,000...but I just had to pay it.” | P30 |

### Negative Experiences with Healthcare Providers

Participants’ negative experiences with healthcare providers often centered on the perception that providers weren’t listening to them or taking their concerns and symptoms seriously. Participants also reported that providers failed to explain the need for medications or procedures, a problem that was especially pronounced for women who had cesareans and felt unprepared for the surgery.

People who had previous pregnancies and experiences with labor and delivery were particularly distressed when their symptoms were dismissed. As one participant relayed: “The doctors just didn’t listen. Nobody was really listening to me, no matter how much I tried to tell them there’s something wrong with me… so that was my frustration and sadness because I felt like I was dying and nobody cared. Nobody cared about my kids. Nobody cared about trying to save me… they just kept brushing everything off” (P05).

A few participants felt their healthcare providers were motivated by financial considerations rather than care and concern for patients: “Yeah, to them it’s just money, insurance, getting patients in and out. But I just had a baby like 48 hours ago, and I’m telling you like something is still not right” (P16).

Several women who participated in this study had unplanned cesarean sections, and they recalled how their questions about the procedure were ignored or they felt forced to have the procedure. As one participant shared: “I usually think the doctor could really do more by looking for alternatives for me before telling me to take the bold step” (P21). They also felt unsupported after giving birth and suggested additional check-ins immediately following the birth. As one participant noted, “You have a cesarean section and you’re in a room by yourself and it’s just like that’s a shock…They just need to have a little bit more there. They always come in and check on the baby. Like you gotta ask mom--is she okay?” (P16).

Analyses of the stories relayed by participants revealed many missed opportunities for providers to establish a sense of caring and connection with patients that would significantly improve the patient experience and, in some cases, may have influenced clinical health outcomes. For example, one participant decided not to take a prescribed medication and, when asked a follow up question by the community researcher about whether she had reached out to the provider said: “[The doctor] knew what she gave me wasn’t going to stop what is, like not stop it totally, but it’s going to suppress the throwing up. But when you give me a medication, I read the good and the bad of it…I’m like it’s not going to kill me, it’s just for nine months” (P23).

Additional quotes on experiences with healthcare providers are available in Table 2.

**Table 2.**
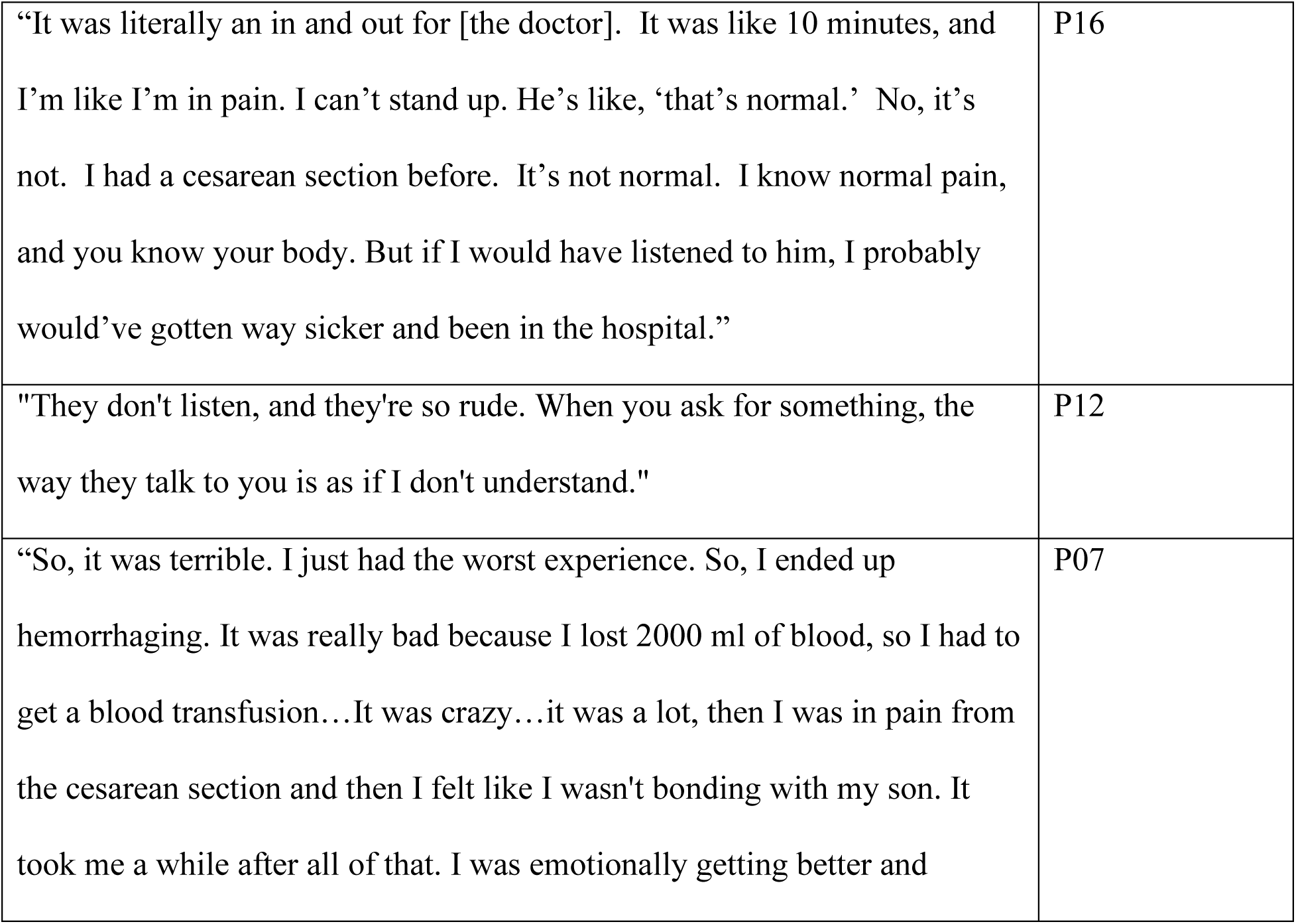
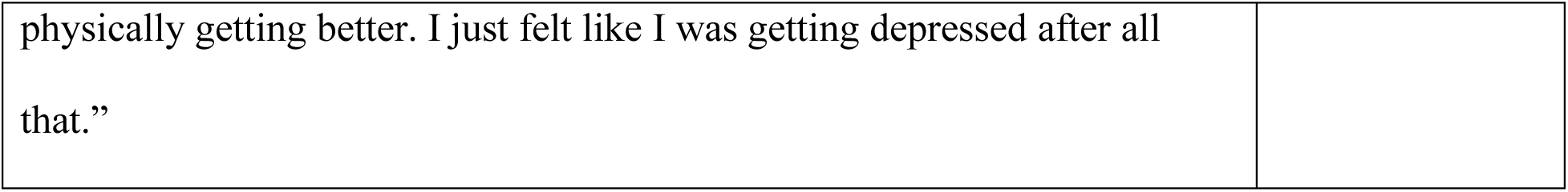
Additional Representative Quotes for Negative Experiences with Healthcare Providers.

|  |  |
| --- | --- |
| “It was literally an in and out for [the doctor]. It was like 10 minutes, and I’m like I’m in pain. I can’t stand up. He’s like, ‘that’s normal.’ No, it’s not. I had a cesarean section before. It’s not normal. I know normal pain, and you know your body. But if I would have listened to him, I probably would’ve gotten way sicker and been in the hospital.” | P16 |
| "They don't listen, and they're so rude. When you ask for something, the way they talk to you is as if I don't understand." | P12 |
| “So, it was terrible. I just had the worst experience. So, I ended up hemorrhaging. It was really bad because I lost 2000 ml of blood, so I had to get a blood transfusion...It was crazy...it was a lot, then I was in pain from the cesarean section and then I felt like I wasn't bonding with my son. It took me a while after all of that. I was emotionally getting better and | P07 |

|  |
| --- |
| physically getting better. I just felt like I was getting depressed after all that.” |

### Preference for Racial Concordance and Experiences of Discrimination

Participants noted that they were more comfortable when their providers were Black or from similar cultural backgrounds and tried to find Black providers. As one participant said, “I did a lot of research before I chose this doctor. I don’t want to sound like racist or biased, but I chose her because she’s African-American. And I chose her because of her history with working with women that were my age having babies. And I just wanted to go to someone that could relate to me” (P01). Participants who had Black providers also reported less discrimination based on race. One person stated, “We have more Black [providers] there, so maybe that’s why I’m not experiencing the discrimination” (P30).

However, participants reported experiencing discrimination based on other aspects of their identity including their insurance type (Medicaid vs. other), age, weight, and national origin. For example, one participant reported: “The only thing I can say that really kind of made me feel like a little sideways is the fact that you know they don’t accommodate the bigger people at doctor’s offices with like scales and blood pressure cuffs. Like everything is normal, you know, regular adult size. There’s nothing else additional that they have for people of my size, like the heart doppler or the blood pressure cuff or the weight scale, it doesn’t accommodate me” (P17).

Another participant commented: “Some of the people that went in there, they got better treatment. They were taken first. I feel like I was definitely treated, because I was a young mother, so they’re like ‘whatever. She doesn’t have any health issues. She’s fine, let’s move on” (P07).

Additional quotes about preference for racial concordance and discrimination are included in Table 3.

**Table 3.** Additional Representative Quotes for Preference for Racial Concordance and Experiences of Discrimination.

|  |  |
| --- | --- |
| “I would say as a woman of color. I'm Latina...I hate to even put it in these terms because I don't even think of color but had I been a Caucasian woman, I think getting an appointment sooner for my mental issues at the time would have been easier had I been Caucasian or perhaps had a different address I think that played a part because it took me a while to find a good provider.” | P27 |
| “The chief medical director, or something that was in charge of me, although I had too many other doctors, she's a Black American. So, it didn't really - so, I had many other doctors that were Blacks. I had some Asians. I had pure Whites, but I didn't really feel any bias. “ | P28 |
| “Yeah, but, you know, when you don't have the insurance. They only give me the charity care or something. Some things you need to get, you don't get them because of your status.” | P15 |

### Mental Health and Social Support

Many participants reported mental health challenges, and these issues took place at every phase of the pregnancy and post-partum periods. Several participants were reluctant to seek mental health services because of the stigma associated with needing help, and they noted that it would have been helpful if they had been screened and referred for help earlier in their pregnancies.

Particularly in the post-partum period, the perception or experience that reporting mental health challenges would interfere with their family’s lives or privacy were barriers to seeking services: “I think they should screen more for women that are going through postpartum, I really think that they should instead of just asking like, ‘hey, how are you feeling, are you ok?’…We’re afraid that if we do say something like, I don’t feel well, I don’t feel like doing this today that our babies are going to be taken” (P09). One participant reported the negative consequences of reporting her post-partum depression: “Now they’re in your business, in your house. They question you… depression is bad, like so bad, but how can we do better without making it seem like you are a criminal? That’s how I felt” (P25).

Many participants wished they had been aware of available resources earlier in their pregnancy, and many were still unaware of available resources at the time of the interview. For example, one participant reflected: “I would say there are a lot of resources here in Newark. They’re just not vocalized enough… There aren’t enough resources to go around for everybody, so we can’t just like, put an advertisement out on TV, but it would be great to know maybe a website or something like, program for parents that could be put on there where parents can actually go and see” (P27).

Lack of childcare for other children and paid family leave for husbands/partners also inhibited participants from accessing care during and after their pregnancies. In contrast, those who did know of services such as medical transportation noted those services as important in helping them receive care and in bolstering their overall satisfaction with their experiences: “So the medical transportation is -- it was great. But I don’t think everyone knows that you can get the medical transportation” (P01).

Additional quotes from participants related to mental health and social support are included in Table 4.

**Table 4.** Additional Representative Quotes for Mental Health and Social Support.

|  |  |
| --- | --- |
| “In the middle of the night when everything is quiet and your mind is just running and racing and thinking, that's when I needed somebody to really talk to.” | P05 |
| “But the mental health support, I think that should be worked in there for women, it should be a natural thing, even if you don't feel like you need it, you don't know if you don't need it until you really hit that wall.” | P09 |
| “So I knew that my water is broken and I had called my spouse at that particular time and I said, I think we need to head over to the hospital, I think my waters have broken. At that point, he was already at work and could not get out. He’s like, I can’t leave work, so you’re going to have to either call an ambulance or call your mom, call somebody, somebody else has got to come and help you.” | P27 |

## Discussion

Very few studies related to Black women’s maternal health have engaged Black women from the community in the design, execution, and analysis of the study. This CBPR study fills a critical gap in the existing literature and reveals actionable insights and possibilities for change.

Structural challenges to obtaining healthcare, including delays and gaps in insurance, uncovered medical costs, long wait times, and accessibility problems when services were not co-located posed barriers to care. Lack of continuous insurance coverage throughout pregnancy was a particular challenge for undocumented immigrants who are only eligible for emergency Medicaid for the period immediately before and after childbirth. Delays in obtaining insurance coverage for pregnant women interfere with prenatal care. For women employed in jobs that do not provide paid sick time, long appointment waiting times pose particular challenges. Even for those with insurance, costly co-pays led people to space out appointments or required them to spend hours negotiating with their insurance companies. Each of these areas could be better addressed by insurers, healthcare organizations, and employers.

This study found a wide range of negative experiences for Black women with healthcare providers including providers not listening, dismissing concerns, and treating women disrespectfully, most of which have also emerged in other studies on this topic.^8–13^ Improving patient experiences is an important goal of healthcare. However, this study also points to how the failure to listen and communicate well with patients can lead to potential negative clinical impacts as evidenced in examples of patients who experienced serious complications after being told their symptoms were normal or decided against taking medications because they were not told of their purpose or did not trust that providers were motivated by caring.

There were numerous missed opportunities for providers to establish trustworthiness, expertise and accessibility, three dimensions which are shown in the literature to make a measurable difference in the quality of the relationship and whether a listener will act on what is said.^28^ Previous work on this topic has shown that for pregnant women of color, having an established relationship with a healthcare provider contributed to feeling cared about and supported; ^29^ however, the structure of care during pregnancy, which frequently includes new healthcare relationships, is often not conducive to establishing ongoing, trusting relationships. The problem was especially pronounced for women having cesarean sections who felt surprised and pushed into the procedure. Educating all pregnant women about the possible reasons for a cesarean section and taking time to explain the reasons for the surgery could improve pregnant women’s experiences. Disparate treatment within the health system requires attention and remediation, as does continued rigorous work to ensure that racism in healthcare is eliminated.

The hospitals that serve Newark, NJ have many healthcare providers and staff at all levels that are Black which allowed pregnant women to find racially concordant providers. Interestingly, most participants in this study did not report feeling discriminated against based on race but did perceive that they received poor treatment based on their insurance type, weight, or age. It is difficult to disentangle whether these participants believed they could not be discriminated against by providers of the same race or whether participants did not experience any discrimination due to race.

Insights gleaned from community-led approaches have the potential to both improve pregnant people’s experiences and their health outcomes. Community researchers with similar backgrounds to the participants were better equipped to ask salient questions, determine the best available research methods, quickly establish trustworthiness and accessibility with participants, and articulate salient solutions. The credibility of community researchers was particularly helpful in discussing stigmatized topics such as mental health. Participants’ reports about their mental health challenges and fears of reporting such challenges may have been shared more readily with community researchers than with traditional researchers.

Moreover, the opportunity to speak one-on-one with a peer and to explore their full pregnancy journey was essential to uncovering information. Indeed, one of the participants said: “Girl, I’m so glad they chose you because sometimes they choose people, because you see the way I’m talking so freely with you is because it’s supposed to be like that… I’m not going to talk to somebody that doesn’t even have kids…So I’m so glad that you actually can relate, and a mother can feel comfortable in talking” (P25). The lived experiences of the community researchers were also important in analyzing the transcripts and determining the key themes of the study.

### Limitations

While multiple strategies were used to inform potential participants about the study, early interviews were drawn from the social networks of the community researchers. Outreach from a broader set of community based organizations did expand the study’s reach but the way the sample was generated limits the generalizability of the findings. In addition, the university’s required online registration system, which is not easy to navigate, may have dissuaded some participants from completing the consent process.

Another study limitation is that the challenges faced in this particular urban area may not be reflective of the challenges faced throughout the country. However, the opportunity to learn from in-depth interviews with Black mothers conducted by Black mothers helps illuminate the many areas that merit attention and, with improvements, could make a real and meaningful difference both clinically and experientially for pregnant people. The findings amplify those of other investigations across the country that focus on improving maternal and birth equity.

## Conclusions

There is a growing awareness of the significant disparities in maternal health outcomes. Most public attention and descriptive studies have focused on maternal mortality and severe maternal morbidity while overlooking the broad experiences women have throughout the pre-natal, labor, delivery, and post-partum period. This investigation, which utilizes CBPR, illuminates a broad range of potential improvements that can be made by multiple sectors which would improve pregnant people’s experiences and outcomes. Further, research which equitably involves community members in every aspect of the investigation has the potential to address many of the concerns related to equity that have been recently raised about research and to contribute new insights and solutions to complex public health problems.

## Data Availability

All data produced in the present study are available upon reasonable request to the authors.

## Acknowledgements

The Greater Newark Health Care Coalition partnered with the university-based research team on recruitment of the community researchers and study participants and is the lead agency on the overall Safer Childbirth Cities of which this study is a component.

